# Persistence of antibody and T cell responses to the Sinopharm/BBIBP-CorV vaccine in Sri Lankan individuals

**DOI:** 10.1101/2021.10.14.21265030

**Authors:** Chandima Jeewandara, Inoka Sepali Aberathna, Pradeep Darshana Pushpakumara, Achala Kamaladasa, Dinuka Guruge, Ayesha Wijesinghe, Banuri Gunasekera, Shyrar Tanussiya, Heshan Kuruppu, Thushali Ranasinghe, Shashika Dayarathne, Osanda Dissanayake, Nayanathara Gamalath, Dinithi Ekanayake, Jeewantha Jayamali, Deshni Jayathilaka, Madushika Dissanayake, Tibutius Thanesh Jayadas, Anushika Mudunkotuwa, Gayasha Somathilake, Michael Harvie, Thashmi Nimasha, Saubhagya Danasekara, Ruwan Wijayamuni, Lisa Schimanski, Pramila Rijal, Tiong K. Tan, Tao Dong, Alain Townsend, Graham S. Ogg, Gathsaurie Neelika Malavige

## Abstract

**Background:** To determine the kinetics and persistence of immune responses following the Sinopharm/BBIBP-CorV, we investigated immune responses in a cohort of Sri Lankan individuals.

**Methods:** SARS-CoV-2 specific total antibodies were measured in 20-to-39 year (n=61), 40-to-59-year and those >60 years of age (n=22) by ELISA, 12 weeks after the second dose of the vaccine. ACE2 receptor blocking antibodies (ACE2R-Ab), antibodies to the receptor binding domain (RBD) of the ancestral virus (WT) and variants of concern, were measured in a sub cohort. T cell responses and memory B cell responses were assessed by ELISpot assays.

**Results:** 193/203 (95.07%) of individuals had detectable SARS-CoV-2 specific total antibodies, while 67/110 (60.9%) had ACE2R-Ab. 14.3% to 16.7% individuals in the 20 to 39 age groups had detectable antibodies to the RBD of the WT and VOC, while the positivity rates of those >60 years of age was <10%. 14/49 (28.6%) had IFN_γ_ ELISpot responses to overlapping peptides of the spike protein, while memory B cell responses were detected in 9/20 to the S1 recombinant protein. The total antibody levels and ACE2R-Ab declined after 2 to 12 weeks from the second dose, while ex vivo T cell responses remained unchanged. The decline in ACE2R-Ab levels was significant among the 40 to 59 (p=0.0007) and ≥60 (p=0.005) age groups.

**Conclusions:** Antibody responses declined in all age groups, especially in those >60 years, while T cell responses persisted. The effect of waning of immunity on hospitalization and severe disease should be assessed by long term efficacy studies.

## Introduction

The Sinopharm/BBIBP-CorV is an inactivated COVID-19 vaccine, which is currently approved in 65 countries ^1^ and it has been the most widely used vaccine in Sri Lanka. The phase 3 clinical trials showed an efficacy of 78.1% against symptomatic illness ^2^, while all individuals were reported to have seroconverted and shown to have developed neutralizing antibodies by 42 days, following the second dose of the vaccine ^3^. We previously reported that 95% of Sri Lankan individuals seroconverted 2 weeks following the second dose, with lower seroconversion rates in older individuals. ACE2 receptor blocking antibodies, measured by the surrogate neutralizing test (sVNT) were found in 81.25% of individuals following both doses of the vaccine, while antibodies to the receptor binding domain was significantly less than those following natural infection ^4^. Although Sinopharm/ BBIBP-CorV is the main vaccine used by many Asian and Middle East countries, there are limited data regarding its efficacy when used in different countries and for different variants. Furthermore, data regarding the persistence of antibody and T cell responses in fully vaccinated individuals are limited.

Neutralizing antibodies and non-neutralizing antibodies specific to the spike protein have shown to gradually decline following the second dose of BNT162b2 (Pfizer–BioNTech) and AZD1222 ^5^, while the mRNA-1273 vaccine was shown to induce stable responses up to six months after the second dose ^6^. Although the quantity of neutralizing antibodies that are needed for protection from severe disease or to prevent breakthrough infection is not known, it has been shown that the efficacy of BNT162b2 (Pfizer–BioNTech) vaccine declines with time ^7^. Booster doses of BNT162b2 (Pfizer–BioNTech) vaccine resulted in a 11.3 fold reduction in acquisition of infection and 19.5 fold reduction in severe disease compared those who did not receive a booster ^8^. However, the WHO recently stated that although there is evidence of declining vaccine efficacy against mild illness, the efficacy of vaccines against hospitalization and severe disease appears to be high and therefore, more emphasis should be placed on vaccinating vulnerable individuals ^9^. Despite the decline in neutralizing antibodies following vaccination and natural infection with time, spike protein specific memory B cells have shown to persist for a longer duration ^10^. Although these long-lived memory cells are thought to give long lasting protection against severe illness, fully vaccinated elderly individuals in Israel appear to have been susceptible to severe disease and thus benefited from a booster dose ^11^. Based on waning of neutralizing antibody responses and efficacy for certain COVID-19 vaccines with time, booster doses are currently been offered to older individuals and individuals in high risk categories in different countries ^12,13^.

Although there are data regarding the duration and persistence of antibody and T cell responses following many COVID-19 vaccines such as BNT162b2 (Pfizer–BioNTech), AZD1222 and mRNA-1273, there are limited data regarding the persistence of immune responses following Sinopharm/BBIBP-CorV. It is crucial that severe disease and hospitalizations due to COVID-19 are curtailed to a level that it no longer becomes a global threat. While vaccinating all individuals worldwide is essential to end the pandemic, it is also important to monitor the duration of immunity and any changes in vaccine efficacy over time. Therefore, we sought to investigate the persistence of antibody and T cell responses in a Sri Lankan cohort to understand the durability of immune responses to this vaccine in different age groups.

## Methods

We previously evaluated immune responses to Sinopharm.BBIBP-CorV in 323 Sri Lankan individuals from Colombo at 4 weeks after the first dose and 2 weeks after the second dose (6 weeks after the first dose)^4^. In order to determine the persistence of antibody and T cell responses following the second dose, blood samples were obtained at 3 months (12 weeks) following the second dose of the vaccine in 203 individuals from this cohort, while those who reported as being PCR positive/diagnosed of COVID-19 or reported symptoms suggestive of COVID-19 such as fever, sore throat, cough and myalgia were excluded from the 3-month analysis. Demographic and the presence of comorbidities such as diabetes, hypertension, cardiovascular disease and chronic kidney disease was determined by a self-administered questionnaire at the time of recruitment from all participants.

Ethics approval was obtained from the Ethics Review Committee of University of Sri Jayewardenepura.

### Detection of SARS-CoV-2 specific total antibodies, ACE2 receptor blocking antibodies and antibodies to the RBD of SARS-CoV-2 VOCs

The Wantai SARS-CoV-2 Ab ELISA (Beijing Wantai Biological Pharmacy Enterprise, China) was used to detect the presence of SARS-COV-2 specific total antibodies (IgM, IgA and IgG), which detects antibodies to the receptor binding domain (RBD) of the spike protein, according to the manufacturer’s instructions. The surrogate virus neutralization test (sVNT) was used to measure ACE2-receptor blocking antibodies and the haemagglutination test (HAT) was used to measure antibodies to the RBD of VOCs in a sub cohort of individuals (n=110). The Svnt was carried out as previously described according to the manufacturer’s instructions (Genscript Biotech, USA) ^14^. This measures the percentage of inhibition of binding of the RBD to recombinant ACE2 and an inhibition percentage ≥ 25% in a sample was considered as positive for ACE2 receptor blocking antibodies ^15^.

The HAT was carried out using the B.1.1.7 (N501Y), B.1.351 (N501Y, E484K, K417N) and B.1.617.2 versions of the IH4-RBD reagents ^16^, which included the relevant amino acid changes introduced by site directed mutagenesis. The assays were carried out and interpreted as previously described and a titre of 1:20 was considered as a positive response ^17,18^. The HAT titration was performed using 7 doubling dilutions of serum from 1:20 to 1:1280, to determine presence of RBD-specific antibodies. The RBD-specific antibody titre for the serum sample was defined by the last well in which the complete absence of “teardrop” formation was observed.

### Ex vivo IFN_γ_ ELISpot assays and B cell ELISpot assays

Ex vivo IFN_γ_ ELISpot assays were carried out using freshly isolated peripheral blood mononuclear cells (PBMC) obtained from 49 individuals in whom we had previously carried out these assays at 4 weeks and 6 weeks. Due to the limitations in the availability of PBMCs to carry out these assays, B cell ELISpots were only done in 28 individuals, in whom ex vivo ELISpot assays were carried out. The details of these assays are described in supplementary methods.

### Statistical analysis

Details of statistical analysis are included in the supplementary methods.

## Results

### SARS-CoV-2 total antibody responses in different age groups

At 3 months (12 weeks) since receiving the 2^nd^ dose 193/203 (95.07%) of individuals had detectable SARS-CoV-2 specific total antibodies. 59/61 (96.72%) between the ages of 20 to 39, 114/120 (95.0%) of those 40 to 59 and 20/22 (90.91%) of those >60 years were seropositive. There was no significant difference (p value=0.06) between the antibody titres (indicated by antibody index) in the three different age groups (20 to 39, 40 to 59 and ≥60 years) (Figure 1A). However, the total antibody levels (indicated by the antibody index) significantly and inversely correlated with age (Spearman’s r=-0.19, p=0.006), (Figure 1B). There was no significant difference (p-value=0.0760) in the seropositivity rates or the antibody levels (indicated by antibody indices) in those with comorbidities, compared to those who did not have comorbidities. Similarly, the seropositivity rates between males and females were also not found to have a statistically significant difference (p-value=0.3319).

**Figure 1:**
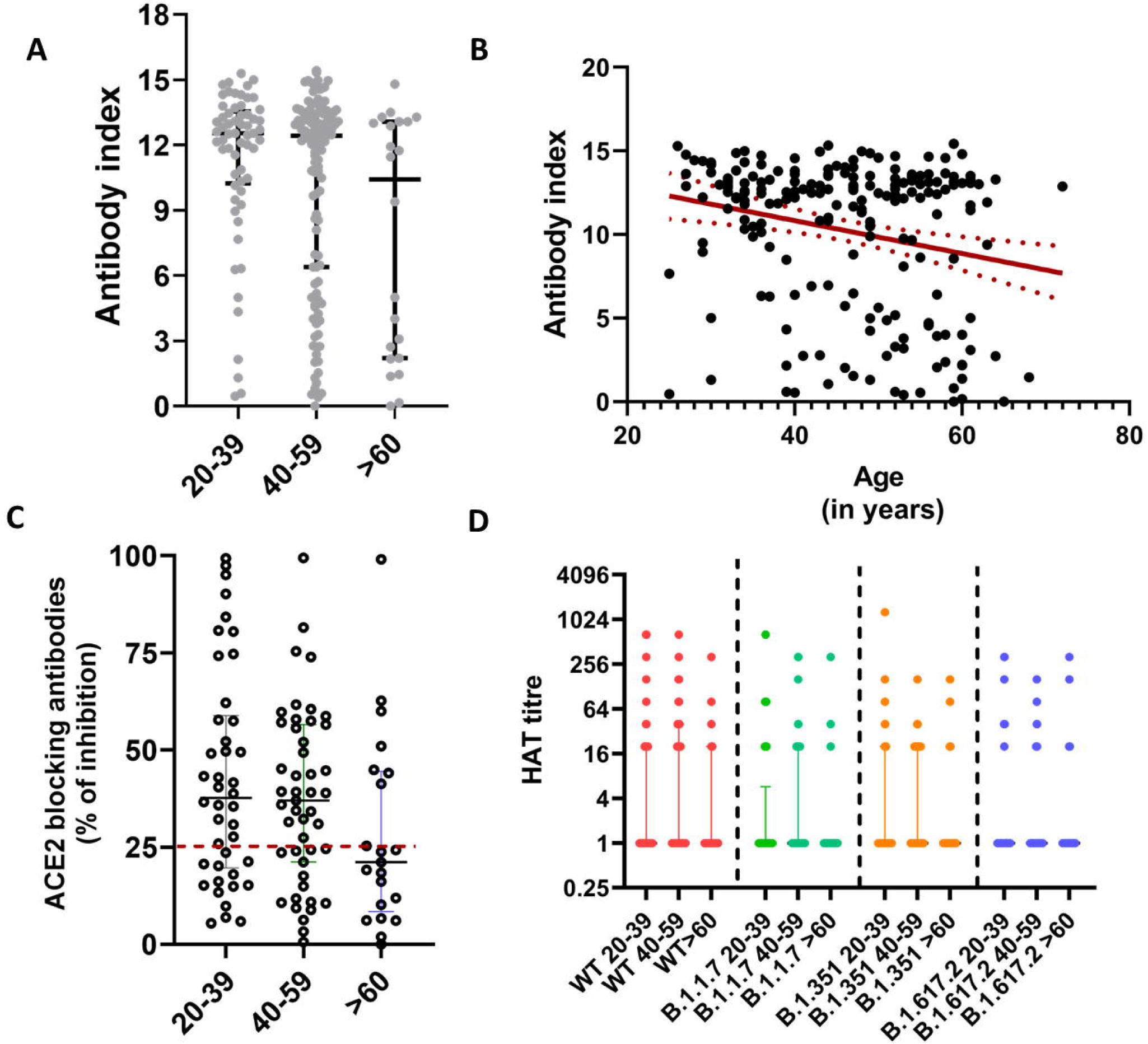
SARS-CoV-2 specific antibody responses 3 months following the second dose (16 week following the first dose) of the Sinopharm/ BBIBP-CorV vaccine. SARS-CoV-2 specific total antibodies were measured in 20-to-39-year old’s (n=61), 40-to-59-year old’s and those >60 years of age (n=22) by ELISA and no significant difference was seen between the age groups (p=0.06) based on the Kruskal-Wallis test (A). The total antibody titres inversely correlated (Spearman’s r=-0.19, p=0.006) with age (B) (the red dotted lines indicate the 95% confidence interval). The ACE2 receptor blocking antibodies were measured by the surrogate virus neutralizing test in 20-to-39-year old’s (n=41), 40-to-59-year old’s (n=48) and >60-year old’s (n=21) and no significant difference was seen (p=0.11) between the age groups based on the Kruskal-Wallis test (C). Antibodies were measured to the receptor binding domain of the ancestral SARS-CoV-2 virus (WT) and to B.1.1.7, B.1.351 and B.1.617.2 using the haemagglutination test (HAT) in 20-to-39-year old’s (n=41), 40-to-59-year old’s (n=48) and >60 year old’s (n=21) and no significant difference was seen (p=0.11) between the age groups based on the Kruskal-Wallis test for different variants (D). All tests were two-tailed. The lines indicate the median and the inter quartile range.

### SARS-CoV-2 specific ACE2-receptor blocking antibodies in different age groups

The surrogate neutralizing antibody test (sVNT) that measures ACE2 receptor blocking antibodies was carried out in a subset of individuals of cohort (n=110) and 67 (60.9%) gave a positive result. 27/41 (65.85%) in the 20 to 39 age group, 32/48 (66.67%) in the 40 to 59 age group and 8/21 (38.10%) of the ≥60-year age group gave a positive result. There was no significant difference (p=0.06) between the ACE2 receptor blocking antibody positivity (% of inhibition >25) in different age groups. There were also no significant differences in the levels of ACE2 receptor blocking antibodies in different age groups (p=0.11) (Figure 1C).

### Hemagglutination test (HAT) to detect antibodies to the receptor binding domain of SARS-CoV-2 and its variants of concern (VOCs)

The HAT assay was carried out to measure positivity rates and the antibody titres to the ancestral strain (WT), and the VOCs B.1.1.7, B.1.351 and B.1.617.2 in the same individuals in whom the sVNT assays were carried out (n=110). The proportion of individuals who gave a positive result for the WT and the different VOCs and their mean, standard deviation, median values with the IQR (HAT titres) are shown in Table 1. As determined by the Friedman test, the HAT titres for the WT were found to be significantly higher than the HAT titres to the different VOCs for the age groups 20 to 39 (p=0.002) and 40 to 59 (p=0.0001). The post-hoc tests for multiple comparisons in the 20 to 39 group showed that the HAT titres for the WT was significantly higher than that for B.1.17 (p=0.004) and B.617.2 (p=0.002). In the 40 to 59 age group, the HAT titres to the WT were significantly higher than for B.1.1.7 (p=0.001), B.1.315 (p=0.006) and B.1.617.2 (p=<0.001). However, the ≥60-year age group had low HAT titres for WT and all VOC, and there were no significant differences (p=0.286) between responses (Figure 1D).

**Table 1:**
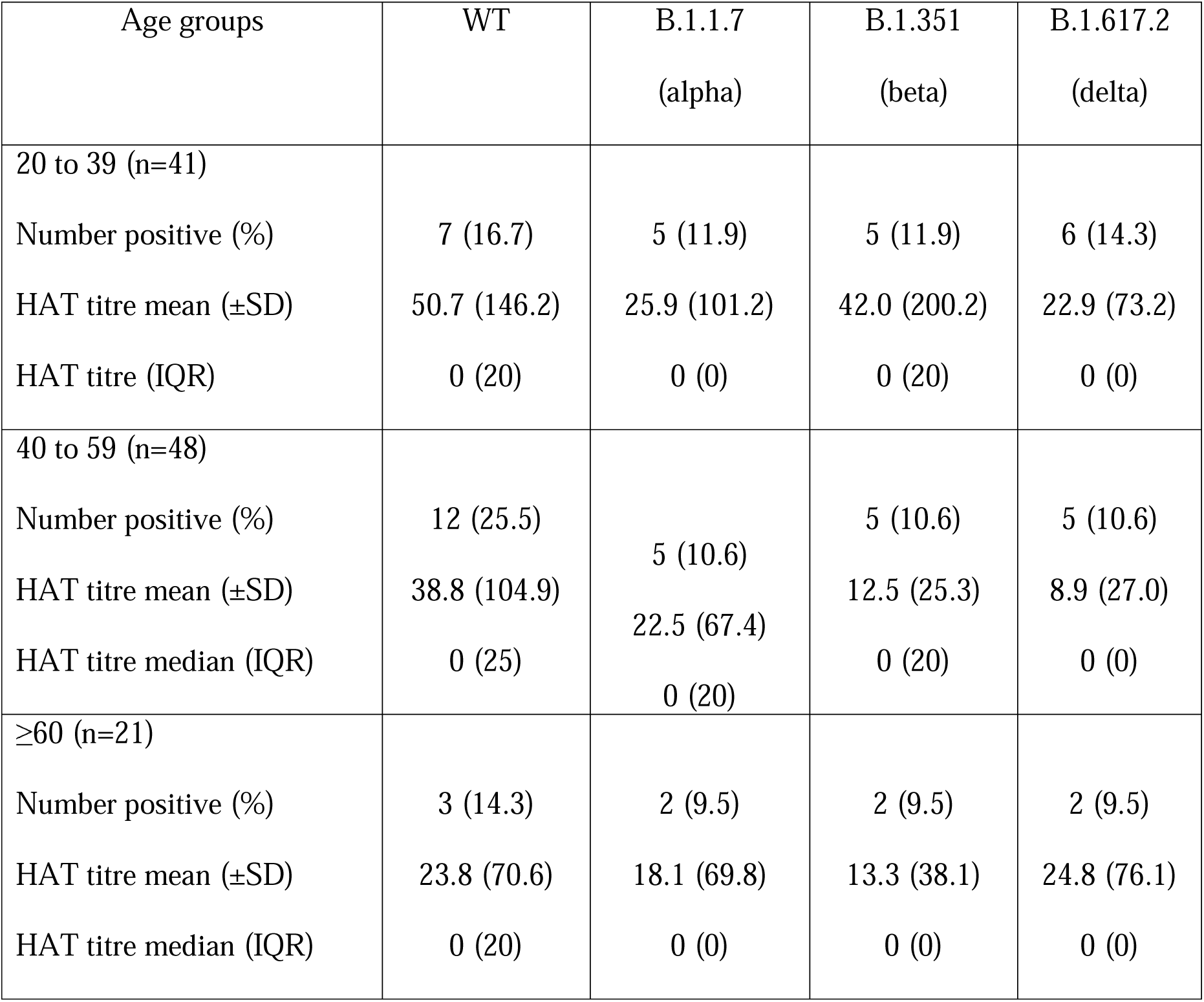
Antibody responses to the receptor binding domain of the ancestral SARS-CoV-2 virus (WT) and to B.1.1.7, B.1.351 and B.1.617.2 using the haemagglutination test (HAT)

| Age groups | WT | B.1.1.7<br>(alpha) | B.1.351<br>(beta) | B.1.617.2<br>(delta) |
| --- | --- | --- | --- | --- |
| 20 to 39 (n=41) |  |  |  |  |
| Number positive (%) | 7 (16.7) | 5 (11.9) | 5 (11.9) | 6 (14.3) |
| HAT titre mean (±SD) | 50.7 (146.2) | 25.9 (101.2) | 42.0 (200.2) | 22.9 (73.2) |
| HAT titre (IQR) | 0 (20) | 0 (0) | 0 (20) | 0 (0) |
| 40 to 59 (n=48) |  |  |  |  |
| Number positive (%) | 12 (25.5) | 5 (10.6) | 5 (10.6) | 5 (10.6) |
| HAT titre mean (±SD) | 38.8 (104.9) | 22.5 (67.4) | 12.5 (25.3) | 8.9 (27.0) |
| HAT titre median (IQR) | 0 (25) | 0 (20) | 0 (20) | 0 (0) |
| $\geq 60$ (n=21) | | | | |
| Number positive (%) | 3 (14.3) | 2 (9.5) | 2 (9.5) | 2 (9.5) |
| HAT titre mean (±SD) | 23.8 (70.6) | 18.1 (69.8) | 13.3 (38.1) | 24.8 (76.1) |
| HAT titre median (IQR) | 0 (20) | 0 (0) | 0 (0) | 0 (0) |

### Ex vivo ELISpot responses in the different age groups

To investigate the T cell responses in these two cohorts, we carried out ex vivo IFN_γ_ ELISpot responses in 49 individuals in different age groups, in those who were recruited by us to study the T cell responses. IFN_γ_ ELISpot responses to S1 overlapping pool of peptides (median 100, IQR 47.5 to 260 SFU/1 million PBMCs) were significantly higher (p=0.008) than those for the S2 pool (median 55, IQR 14 to 190 SFU/1 million PBMCs). The threshold for a positive response was set at 234 SFU/1 million PBMCs and accordingly, 14/49 (28.6%) of the individuals gave a positive response. There was no significant difference (p=0.33) in the ex vivo ELISpot responses between the different age groups (Figure 2A). 7/16 (43.6%) of those between 20 to 39 years of age, 3/21 (14.3%) of those between 40 to 59 years of age and 4/12 (33.3%) had a positive response to S1 pool of peptides. There was no correlation between the age of the individual and ex vivo IFN_γ_ ELISpot responses to either S1 (Spearman’s r=-0.22, p=0.12) or S2 (Spearman’s r=-0.19, p=0.18) pool of peptides. The ex vivo overall IFN_γ_ ELISpot responses significantly correlated with the SARS-CoV-2 specific total antibodies (Spearman’s r=0.33, p=0.02), but not with ACE2 receptor blocking antibodies (Spearman’s r=0.20, p=0.16).

**Figure 2:**
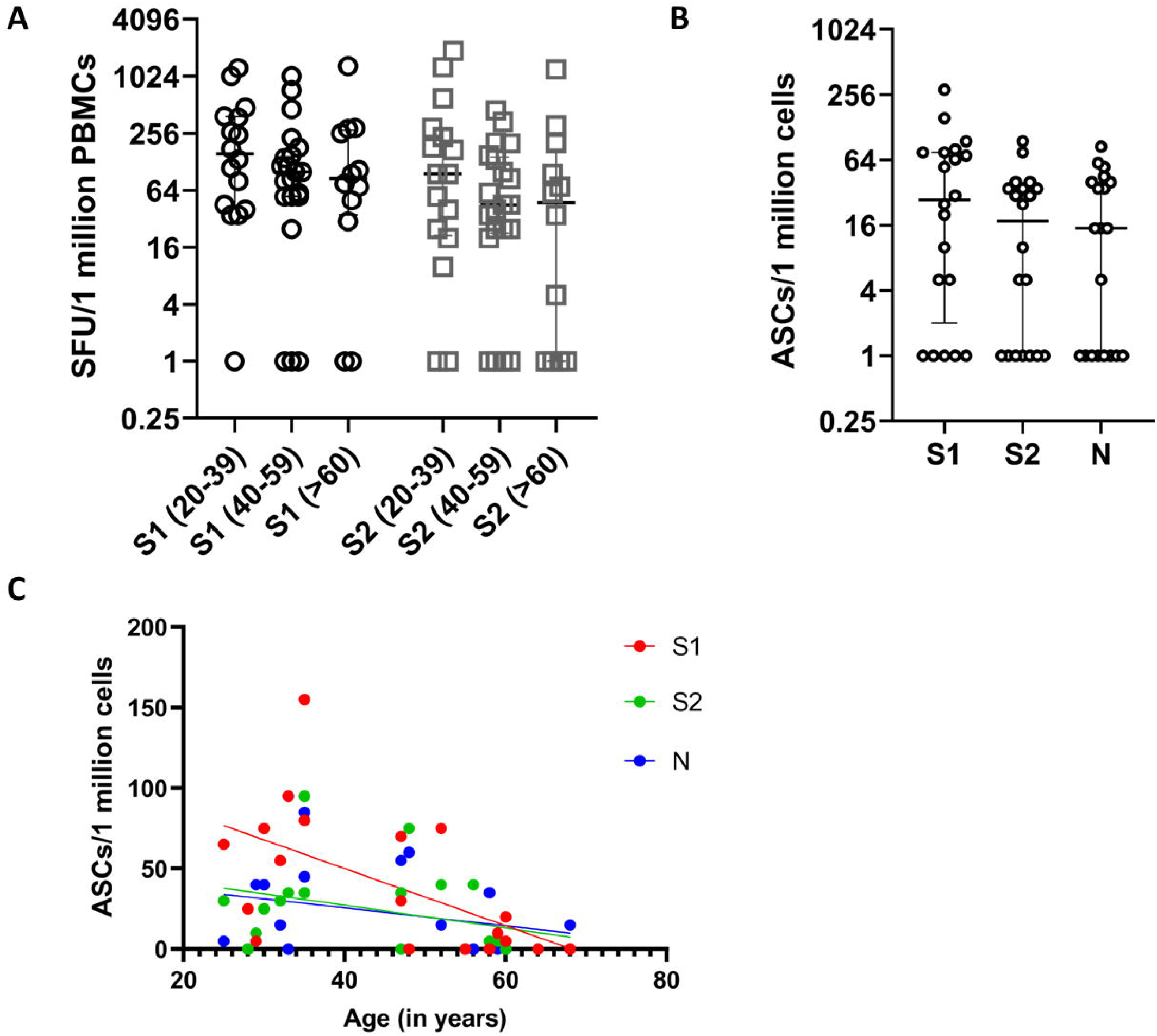
Ex vivo IFN_γ_ ELISpots responses and B cell ELISpot responses 3 months following the second dose (16 week following the first dose) of the Sinopharm/ BBIBP-CorV vaccine. Ex vivo IFN_γ_ ELISpots responses were measured to the S1 and S2 overlapping pool of peptides in 49 individuals between the age groups of 20 to 39 (n=16), 40 to 59 (n=21) and in >60 year olds (n=12). There was no significant difference (p=0.33) in the ex vivo ELISpot responses between the different age groups based on the Kruskal-Wallis test (A). The frequency of antibody secreting cells (ASCs) to S1, S2 and N recombinant proteins were assessed by B cell ELISpot assays in 20 individuals. Using the Kruskal-Wallis test, we found that there was no difference between the frequency of ASCs for S1, S2 and the N protein in different individuals (B). The frequency of ASCs to the S1 protein significantly decreased with age (Spearman’s r=- 0.57, p=0.01), but not for S2 (Spearman’s r=0.37, p=0.12) or N (Spearman’s r=0.34, p=0.14) (C).

### The frequency of antibody secreting cells (ASCs) in those who received Sinopharm.BBIBP-CorV

B cell ELISpot assays for S1, S2 and N recombinant proteins were carried out in 20/49 individuals, who were also included for evaluating of ex vivo T cell responses. The threshold of a positive response was set at 44.1 ASCs/1 million cells for S1, 32.9 for S2 and 19.1 for the N protein. 9/20 (45%) individuals gave a positive response for S1, 7/20 (35%) individuals responded to S2 and, 8/20 (40%) for the N protein. Using the Friedman test, we found that there was no difference between the frequency of ASCs for S1, S2 and the N protein in different individuals (Fig 2B). Although the frequency of ASCs to the S1 protein significantly decreased with age (Spearman’s r=-0.57, p=0.01), no such association was seen with responses to S2 and the N protein (Fig 2C).

### Kinetics of antibody responses and T cell responses over time

Although initially we planned to follow the whole cohort that was initially recruited to the study, at the time of recruitment, we could only obtain blood samples from 174 individuals at all 4 time points (baseline, 4 weeks from first dose, 2 weeks from second dose and 3 months from second dose) to measure SARS-CoV-2 specific total antibodies. For this analysis, we were able to follow 49 individuals in the 20 to 39 age group, 108 in the 40 to 59 age group and 17 in ≥60 years age group. From the second dose, the SARS-CoV-2 total antibodies, measured by the Wantai antibody assay, declined in all the age groups but this decline was not significant in any of the age groups by the Friedman test (Figure 3A).

**Figure 3:**
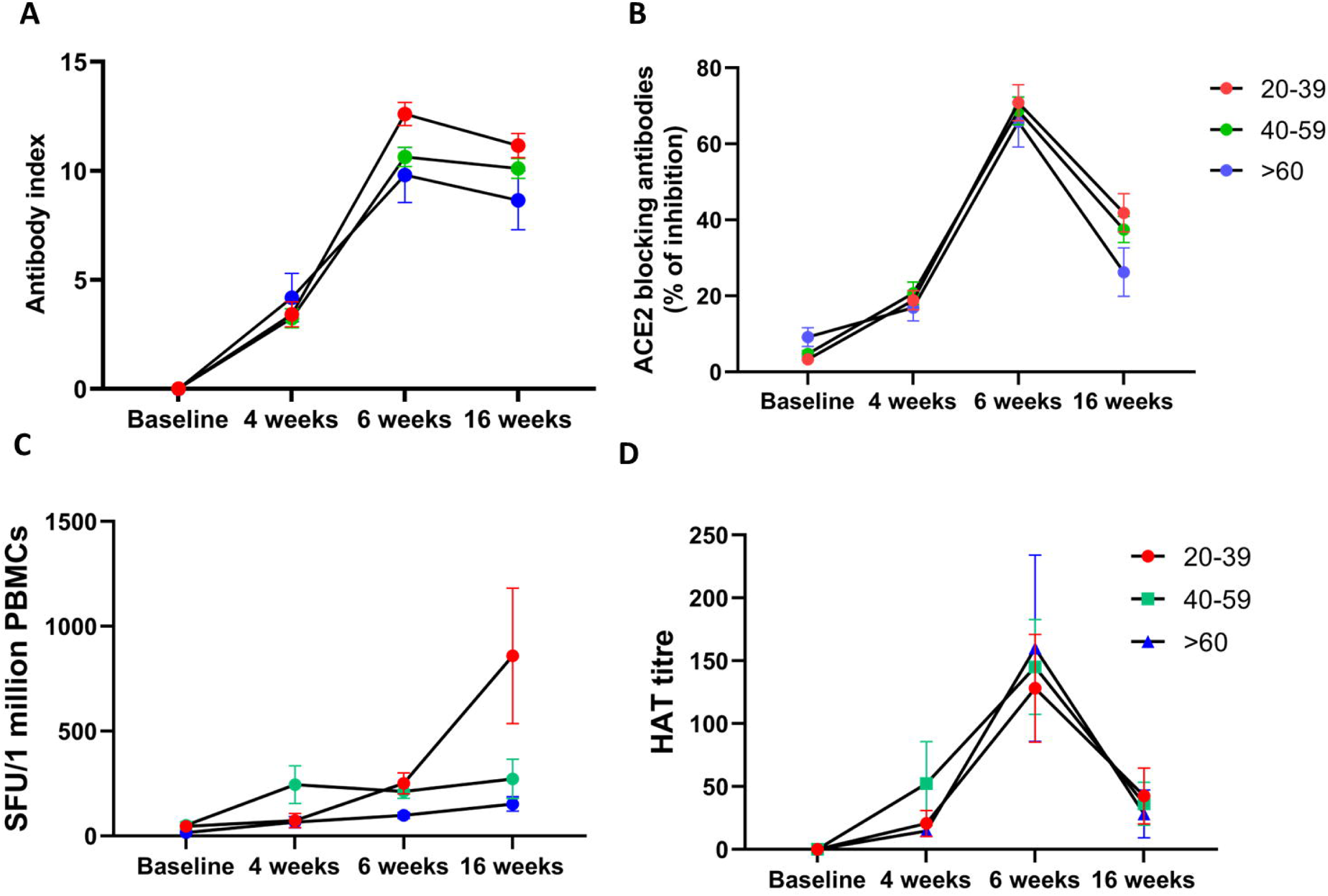
Kinetics of antibody and T cell responses over time. SARS-CoV-2 total antibodies were measured in 174 individuals, (49 in 20 to 39, 108 in the 40 to 59 and 17 in ≥60 years age group), at baseline, 4 weeks after the first dose, 6 weeks (2 weeks after the 2^nd^ dose) and at 16 weeks (12 weeks/3 months) after the second dose by ELISA. The decline in antibody responses from 6 to 16 weeks was not statistically significant in any age group (A). ACE2 receptor antibodies were measured by the surrogate virus neutralizing test in 92 individuals (32 in 20 to 39, 43 in 40 to 59 and 17 in the ≥60 years age group). The decline in antibody levels from 6 to 16 weeks was significant in 40 to 59 (p=0.0007) and ≥60 (p=0.005) age group (B). Ex vivo IFN_γ_ ELISpot responses to the S protein overlapping pool of peptides were measured in 37 individuals, with 10 in the 20 to 39 age group, 18 in the 40 to 59 age group and 9 in the >60 age group. There was no difference in responses between 6 to 16 weeks. Antibodies to the RBD of the WT was measured by the haemagglutination assay test in 92 individuals (32 in 20 to 39, 43 in 40 to 59 and 17 in the ≥60 years age group). The decline in antibody levels from 6 to 16 weeks was significant in 20 to 39 (p<0.0001), 40 to 59 (p<0.0001) and ≥60 (p=0.0002) age group (D). The lines indicate the mean and the error bars the standard error of the mean. All tests were two-tailed.

We could only recruit 92 individuals to measure the variability of ACE2 receptor blocking antibodies over time. These assays were carried out in 32 individuals in the 20 to 39 age group, 43 in the 40 to 59 age group and 17 in the ≥60 years age group. We saw a decline in ACE2 receptor blocking antibodies in all age groups from the 2^nd^ dose, which was more marked in the ≥60 year old age group (Figure 3B). The decline in antibody levels were significant among the 40 to 59 (p=0.0007) and ≥60 (p=0.005) age groups. There was no significant difference in ACE2 receptor blocking antibodies for 4 weeks after the first dose, compared to 3 months after the second dose in 20 to 39 year olds (p=0.09), 40 to 59 year olds (p=0.22) and >60 year olds (p>0.99). In the 20 to 39 age group, there was a 1.7 fold reduction in the levels of ACE2 receptor blocking antibodies (% of inhibition), while in the 40 to 59 age group there was a 1.8 fold reduction and a 2.5 fold reduction in the >60 year age group.

To explore the kinetics of ex vivo IFN_γ_ ELISpot responses over time, we only had all four time points in 37 individuals, with 10 in the 20 to 39 age group, 18 in the 40 to 59 age group and 9 in the >60 age group. In contrast to observations with the antibody responses, we saw the frequency of responses to the overlapping peptides of the spike protein increase over time in those in the 20 to 39 age group, while the responses in the 40 to 59 and >60 age group remained unchanged (Figure 3C). However, the increase in the ex vivo IFN_γ_ responses in the 20 to 39 group from 6 weeks to 3 months (16 weeks), was not significant (p>0.99).

We also investigated the change in the antibody titres to the RBD of the WT in 92 individuals (32 in 20 to 39, 43 in 40 to 59 and 17 in the ≥60 years age group) by the haemagglutination assay. The decline in antibody levels from 6 to 16 weeks was significant in 20 to 39 (p<0.0001), 40 to 59 (p<0.0001) and ≥60 (p=0.0002) age group (D). The HAT titres were only significantly higher than the 4 week values (4 weeks after a single dose) in the 20 to 39 age group (p=0.003), whereas the HAT titre to the RBD of the WT was similar the responses seen at 4 weeks in the 40 to 59 (p>0.99) and >60 age group (p>0.99).

## Discussion

In this study, we have determined the antibody and T cell responses in a cohort of Sinopharm/BBIBP-CorV vaccinated individuals that we have been following throughout for 16 weeks. 3 months (12 weeks) following the second dose we found that 95.07% of individuals had detectable SARS-CoV-2 specific total antibodies, although the antibody levels significantly declined with age. Although we did not measure neutralizing antibodies, we used the sVNT to measure ACE2 receptor blocking antibodies, which have shown to correlate with neutralizing antibodies ^14^. Based on this assay, 60.09% of individuals had ACE2 receptor blocking antibodies, although only 38.1% of those >60 years of age had detectable blocking antibodies. These ACE2 receptor blocking antibodies had declined in all individuals within a period of 16 weeks from receiving the second dose. Significant reductions from 2 weeks from the second dose was seen in individuals >40 years of age. Although the clinical implications of the decline in these antibody responses are not known, neutralizing antibodies have shown to correlate with protection from infection ^19^. Although neutralizing antibodies also are shown to correlate with prevention of symptomatic infection^20^, it is yet unclear if it prevents severe illness.

Using the HAT assay, we measured antibodies to the RBD of the WT and VOCs in these different age groups. 14.3% to 16.7% individuals in the 20 to 39 age groups had detectable antibodies by this assay, while the positivity rates of those >60 years of age was <10%. Interestingly, significant differences were not seen between positivity rates to WT vs VOCs in these individuals, although the mean HAT titres were lower in all individuals to B.1.351 compared to other VOCs. The HAT assay was also shown to correlate with neutralizing antibodies ^21^. Therefore, based on the HAT assay and the sVNT assay, the presence of neutralizing antibodies levels appears to be low or undetectable in all age groups, but especially in those >60 years of age. At 12 weeks from the second dose a 1.7 to 2.5 reduction of ACE2 receptor blocking antibodies were seen, with significant reductions in HAT titres to the RBD of the WT in all age groups. It was shown that the BNT162b2 (Pfizer–BioNTech) and AZD1222 too had a two-fold and five-fold reduction of antibodies to the S-protein respectively, 70 days following the second dose of the vaccine ^5^. Our data showed that the ACE2 receptor blocking antibody levels were detectable in 75.9% individuals at >16 weeks after a single dose of AZD1222 ^22^, whereas only 60.09% of individuals who received the two doses of Sinopharm/BBIBP-CorV vaccine were positive for these antibodies at 12 weeks following the second dose. Therefore, although a significant decline has also been observed with BNT162b2 (Pfizer–BioNTech) and AZD1222 with time, it would be important to compare the decline in antibody responses in different vaccines with time. However, as it is unclear if the reduction in neutralizing antibodies we have detected in the circulation would result in increased susceptibility to severe disease. Long-term efficacy studies are urgently needed to determine if such a reduction in circulating antibodies, while B and T cell memory is maintained, will result in enhanced risk of severe outcomes from infection.

Memory B cell responses have shown to be more durable and have shown to provide long lasting immunity ^10^ and have shown to increase with time following natural infection ^23^. We found that 40 to 45% of individuals had SARS-CoV-2 specific ASCs, 12 weeks following the second dose of the vaccine, while 28.6% of individuals had detectable ex vivo T cell responses. Although the SARS-CoV-2 specific antibody responses had declined with time, the frequency of ex vivo IFN_γ_ ELISpot responses increased in the 20 to 39 age group from 2 weeks following the second dose to 12 weeks, while it remained unchanged in those in the 40 to 59 and >60 age group. Early appearance of T cell responses have been shown to associate with reduced clinical disease severity ^24,25^. Therefore, although antibody responses declined over time in the vaccinees, the presence of a sustained memory B cell and a T cell response, could prevent the occurrence of severe illness, although breakthrough infection might still occur. Due to the limited number of B cell ELISpots carried out, we did not have sufficient number of individuals to compare the variation of ASCs over time.

In summary, we have described the immune responses to the Sinopharm/BBIBP-CorV vaccine, 12 weeks following the second dose of the vaccine. We show that while the SARS-CoV-2 specific total antibodies, and especially ACE2 receptor blocking antibodies and antibodies to the RBD significantly decline, the memory T cell and B cell responses persisted. Since the ACE2 receptor blocking antibodies was shown to significantly decline in all age groups and especially in the elderly, it is important to carry out long term efficacy studies to assess the waning of immunity on hospitalization and severe disease in order to decide on booster doses in different populations.

## Data Availability

All data produced in the present work are contained in the manuscript.

## Acknowledgement

We are grateful to the Allergy, Immunology and Cell Biology Unit, University of Sri Jayewardenepura; the NIH, USA (grant number 5U01AI151788-02), UK Medical Research Council and the Foreign and Commonwealth Office for support. T.K.T. is funded by the Townsend-Jeantet Charitable Trust (charity number 1011770) and the EPA Cephalosporin Early Career Researcher Fund. A.T. are funded by the Chinese Academy of Medical Sciences (CAMS) Innovation Fund for Medical Science (CIFMS), China (grant no. 2018-I2M-2-002).

## Reference

1. Team MCVT. COVID-19 vaccine tracker. McGill COVID19 Vaccine Tracker Team. (https://covid19.trackvaccines.org/vaccines/approved/).

2. Al Kaabi N, Zhang Y, Xia S, et al. Effect of 2 Inactivated SARS-CoV-2 Vaccines on Symptomatic COVID-19 Infection in Adults: A Randomized Clinical Trial. JAMA : the journal of the American Medical Association 2021;326(1):35–45. DOI: 10.1001/jama.2021.8565.

3. Xia S, Zhang Y, Wang Y, et al. Safety and immunogenicity of an inactivated SARS-CoV-2 vaccine, BBIBP-CorV: a randomised, double-blind, placebo-controlled, phase 1/2 trial. The Lancet infectious diseases 2021;21(1):39–51. DOI: 10.1016/S1473-3099(20)30831-8.

4. Jeewandara C, Aberathna IS, Pushpakumara PD, et al. Antibody and T cell responses to Sinopharm/BBIBP-CorV in naïve and previously infected individuals in Sri Lanka. medRxiv 2021:2021.07.15.21260621. DOI: 10.1101/2021.07.15.21260621.

5. Shrotri M, Navaratnam AMD, Nguyen V, et al. Spike-antibody waning after second dose of BNT162b2 or ChAdOx1. Lancet 2021;398(10298):385–387. DOI: 10.1016/S0140-6736(21)01642-1.

6. Doria-Rose N, Suthar MS, Makowski M, et al. Antibody Persistence through 6 Months after the Second Dose of mRNA-1273 Vaccine for Covid-19. The New England journal of medicine 2021;384(23):2259–2261. DOI: 10.1056/NEJMc2103916.

7. Thomas SJ, Moreira ED, Jr., Kitchin N, et al. Safety and Efficacy of the BNT162b2 mRNA Covid-19 Vaccine through 6 Months. The New England journal of medicine 2021. DOI: 10.1056/NEJMoa2110345.

8. Bar-On YM, Goldberg Y, Mandel M, et al. Protection of BNT162b2 Vaccine Booster against Covid-19 in Israel. The New England journal of medicine 2021;385(15):1393–1400. DOI: 10.1056/NEJMoa2114255.

9. WHO. Interim statement on booster doses for COVID-19 vaccination. WHO: 4th October 2021 2021. (https://www.who.int/news/item/04-10-2021-interim-statement-on-booster-doses-for-covid-19-vaccination).

10. Turner JS, Kim W, Kalaidina E, et al. SARS-CoV-2 infection induces long-lived bone marrow plasma cells in humans. Nature 2021;595(7867):421–425. DOI: 10.1038/s41586-021-03647-4.

11. Dolgin E. COVID vaccine immunity is waning - how much does that matter? Nature 2021;597(7878):606–607. DOI: 10.1038/d41586-021-02532-4.

12. Prevention CfDCa. CDC Statement on ACIP Booster Recommendations. Center for Disease Control and Prevention, September 24, 2021 2021. (https://www.cdc.gov/media/releases/2021/p0924-booster-recommendations-.html).

13. Immunisation TJCoVa. JCVI statement regarding a COVID-19 booster vaccine programme for winter 2021 to 2022. The Joint Committee on Vaccination and Immunisation 14th September 2021 2021. (https://www.gov.uk/government/publications/jcvi-statement-september-2021-covid-19-booster-vaccine-programme-for-winter-2021-to-2022/jcvi-statement-regarding-a-covid-19-booster-vaccine-programme-for-winter-2021-to-2022).

14. Tan CW, Chia WN, Qin X, et al. A SARS-CoV-2 surrogate virus neutralization test based on antibody-mediated blockage of ACE2-spike protein-protein interaction. Nature biotechnology 2020;38(9):1073–1078. DOI: 10.1038/s41587-020-0631-z.

15. Jeewandara C, Jayathilaka D, Gomes L, et al. SARS-CoV-2 neutralizing antibodies in patients with varying severity of acute COVID-19 illness. Sci Rep 2021;11(1):2062. DOI: 10.1038/s41598-021-81629-2.

16. Townsend A, Rijal P, Xiao J, et al. A haemagglutination test for rapid detection of antibodies to SARS-CoV-2. Nature Communications 2020:2020.10.02.20205831. DOI: 10.1101/2020.10.02.20205831.

17. Jeewandara C, Kamaladasa A, Pushpakumara PD, et al. Immune responses to a single dose of the AZD1222/Covishield vaccine in health care workers. Nat Commun 2021;12(1):4617. DOI: 10.1038/s41467-021-24579-7.

18. Kamaladasa A, Gunasekara B, Jeewandara C, et al. Comparison of two assays to detect IgG antibodies to the receptor binding domain of the SARSCoV2 as a surrogate marker for assessing neutralizing antibodies in COVID-19 patients. Int J Infect Dis 2021. DOI: 10.1016/j.ijid.2021.06.031.

19. Addetia A, Crawford KHD, Dingens A, et al. Neutralizing Antibodies Correlate with Protection from SARS-CoV-2 in Humans during a Fishery Vessel Outbreak with a High Attack Rate. Journal of clinical microbiology 2020;58(11). DOI: 10.1128/JCM.02107-20.

20. Khoury DS, Cromer D, Reynaldi A, et al. Neutralizing antibody levels are highly predictive of immune protection from symptomatic SARS-CoV-2 infection. Nature medicine 2021;27(7):1205–1211. DOI: 10.1038/s41591-021-01377-8.

21. Lamikanra A, Nguyen D, Simmonds P, et al. Comparability of six different immunoassays measuring SARS-CoV-2 antibodies with neutralizing antibody levels in convalescent plasma: From utility to prediction. Transfusion 2021. DOI: 10.1111/trf.16600.

22. Chandima Jeewandara DG, Pradeep Darshana Pushpakumara, Achala Kamaladasa, Inoka Sepali Aberathna, Shyrar Tanussiya, B Banuri Gunasekera, Ayesha Wijesinghe, Osanda Dissanayake, Heshan Kuruppu, Thushali Ranasinghe, Deshni Jayathilaka, Shashika Dayarathna, Dinithi Ekanayake, MPDJ Jayamali, Nayanathara Gamalath, Anushika Mudumkotuwa, Gayasha Somathilake, Madhushika Dissanayake, Michael Harvie, Thashmi Nimasha, Deshan Madusanka, Tibutius Jayadas, Ruwan Wijayamuni, Lisa Schimanski, Pramila Rijal, Tiong.K. Tan, View ORCID ProfileAlain Townsend, Graham S. Ogg, Gathsaurie Neelika Malavige. Immune responses to a single dose of the AZD1222/Covishield vaccine at 16 weeks in individuals in Sri Lanka. Journal of Immunology (accepted) 2021.

23. Dan JM, Mateus J, Kato Y, et al. Immunological memory to SARS-CoV-2 assessed for up to 8 months after infection. Science 2021;371(6529). DOI: 10.1126/science.abf4063.

24. Lafon E, Diem G, Witting C, et al. Potent SARS-CoV-2-Specific T Cell Immunity and Low Anaphylatoxin Levels Correlate With Mild Disease Progression in COVID-19 Patients. Frontiers in immunology 2021;12:684014. DOI: 10.3389/fimmu.2021.684014.

25. Tan AT, Linster M, Tan CW, et al. Early induction of functional SARS-CoV-2-specific T cells associates with rapid viral clearance and mild disease in COVID-19 patients. Cell reports 2021;34(6):108728. DOI: 10.1016/j.celrep.2021.108728.

