## supplementary figures for "Persistence of antibody and T cell responses to the Sinopharm/BBIBP-CorV vaccine in Sri Lankan individuals"

**Supplementary methods**

Ex vivo IFNγ ELISpot assays

Ex vivo IFNγ ELISpot assays were carried out using freshly isolated peripheral blood mononuclear cells (PBMC). Two pools of overlapping peptides named S1 (peptide 1 to 130) and S2 (peptide 131 to 253) covering the whole spike protein (253 overlapping peptides) were added at a final concentration of 10 µM and incubated overnight ^1,2^. All experiments were done in duplicate and PHA was included as a positive control while media alone was used as a negative control. Briefly, ELISpot plates (Millipore Corp., Bedford, USA) coated with anti-human IFNγ antibody overnight (Mabtech, Sweden), were incubated overnight at 37°C and 5% CO_2_ at a concentration of 100,000 cells/ well. The plates developed with a second biotinylated antibody to human IFNγ and subsequently developed with streptavidin-alkaline phosphatase (Mabtech AB) and colorimetric substrate. The spots were enumerated using an automated ELISpot reader (AID Germany). Background (PBMCs plus media alone) was subtracted and data expressed as number of spot-forming units (SFU) per 10^6^ PBMCs. A positive response was defined as mean±2 SD of the background responses.

B cell ELISpot assays

Briefly, freshly isolated PBMCs were stimulated in a 24 well plate using IL-2 and R848 (a TLR 7/8 agonist) in RPMI supplemented with 10% fetal bovine serum, 1% penicillin streptomycin and 1% glutamine at 4 million cells/well and incubated at 37 °C with 5% CO_2_ for 3 days. They were then washed and rested overnight and 100,000 cells/well were added. 50,000 cells/well were added to the positive control wells. A Human IgG ELISpot kit (Mabtech 3850-2A) was used according to the manufacturer’s instructions to quantify IgG-secreting cells specific to SARS-COV2 S1, S2 and N recombinant proteins, which were coated at 2µg/ml in phosphate buffered saline (PBS). All experiments were carried out in duplicate and anti-human IgG monoclonal capture antibodies, was used as a positive control, and media alone as a negative control. A positive response was defined as mean±2 SD of the background responses. The spots were enumerated using an automated ELISpot reader (AID Germany).

Statistical analysis

The 95% confidence intervals for seropositivity for each age category were calculated using the R software (version 4.0.3) and R-studio (version 1.4.1106). Non-parametric tests such as Mann-Whitney and Kruskal-Wallis were performed at a confidence level of 95% to identify the statistically significant relationships between the age categories and the sex of the individuals with seropositivity and the levels of antibodies. Spearman’s correlation coefficient was used to determine the correlation between antibody, T cell responses and the age of an individual. Moreover, Friedman Tests were performed to identify if there is any significant differences between the antibody levels of the participants with the four different sample collection time points. Similar analyses were carried out for sVNT and HAT results to check if there are any statistically significant differences among the time points.

**References**

1.	Malavige GN, Jones L, Kamaladasa SD, et al. Viral load, clinical disease severity and cellular immune responses in primary varicella zoster virus infection in Sri Lanka. PloS one 2008;3(11):e3789. (In eng) (<http://www.ncbi.nlm.nih.gov/entrez/query.fcgi?cmd=Retrieve&db=PubMed&dopt=Citation&list_uids=19023425> ).

2.	Peng Y, Mentzer AJ, Liu G, et al. Broad and strong memory CD4(+) and CD8(+) T cells induced by SARS-CoV-2 in UK convalescent individuals following COVID-19. Nature immunology 2020;21(11):1336-1345. DOI: 10.1038/s41590-020-0782-6.
